# Age specific COVID-19 undercount in Spain from the regional breakdown of 52–week accumulated mortality rates

**DOI:** 10.1101/2021.03.15.21253667

**Authors:** José María Martín-Olalla

## Abstract

Spanish NUTS3 region records of age specific weekly deaths since the year 2020, records of age specific COVID–19 deaths and age specific population since the year 2020 are used to estimate first age specific all–cause death rates and age specific COVID–19 undercount.

Results shows a statistically significant impact of the pandemic in excess deaths for population aged 40 and elder. Statistically significant COVID–19 undercount is identified only for population aged 80 and elder. Very likely this is the result of the impact of the pandemic in institutionalized population at the early stages of the pandemic.

## 1. INTRODUCTION

The illness designated COVID–19 caused by the severe acute respiratory syndrome coronavirus 2 (SARS-CoV-2) caught worldwide attention since its identification in late 2019[1]. Spain is one of the European countries most impacted by the disease during the 2020–2021 season, mainly during the spring of 2020 at the early stages of the pandemic.

Pandemic numbers —cases and deaths— were quickly tracked by institutions like the John Hopskins Univesity of Medicine https://coronavirus.jhu.edu/map.html. Later, governamental statistics offices provides data on all–cause deaths which showed an huge excursion in many countries[4]. All–cause deaths allow a better understanding of the true impact of the pandemic by including many societal issues.

All–cause excess deaths are expected to differ from COVID–19 deaths due to a myriad of things. To number a few, COVID–19 cases were difficult to identify at the early stages of the pandemic due to lack of testing capabilities and also due to the overwhelming number of infected people which later could be traced by seroprevalence[2] studies and by all–cause death rates[3]; second, the stress put on the public health system may make other causes of death climb. A relative COVID–19 undercount is then defined as the ratio of all–cause excess death rate to COVID–19 death rate. It is a sensitive metric to report worldwide to assess different country capabilities.

In this work, age specific COVID–19 undercount is reported in Spain for a 52–week (year on year, or 364 days) time window. The understanding of age specific death rates is of the utmost importance[5; 6; 7] when addressing the impact of a pandemic like COVID–19.

Results suggest that undercount is statistically significant only for population ages 80 years or older. This age group dominates COVID–19 death toll.

## 2. METHODS AND DATASETS

Confirmed COVID–19 cases and deaths are numbered in Spain by the Centro Nacional de Epidemiología (CNE), equivalent to the CDC after reports from local authorities. Total deaths are numbered by the National Statistics Institute (INE). CNE reports daily cases and deaths on a daily basis. INE reports weekly deaths on a fortnight basis.

INE reports to Eurostat from which the file demo r mwk3 10.tsv can be obtained. The data sets collects weekly deaths disaggregated by region down to NUTS3 level (*N* = 52 in Spain), gender and AGEGRP10 —population grouped in ten-year width bins. The Spanish record runs from 2000 until 2021 week 08 (sunday February 28, 2021) as of March 09, 2021. The record will be updated around March 24th, 2021. The record follows the international week year and accumulates 1104 weeks by now.

The CNE releases the file https://cnecovid.isciii.es/covid19/resources/casos_hosp_uci_def_sexo_edad_provres.csv which reports number of deaths from identified COVID–19 cases by date of death. This record runs from Jan 1st, 2020 to March 15, 2021 and provides provides disaggregation by sex, gender and and province (NUTS3 region) of residence. Unluckily the available gender breakdown is of very poor quality, with a significant fraction of deaths lacking gender assignment.

Finally, the table 31304 https://www.ine.es/jaxiT3/Tabla.htm?t=31304 from INE reports Spanish population at January 1st and July 1st per gender, one-year age group and NUTS3 region since 1970. This record was last updated on early 2021 with population until July 1st, 2020. Population numbers for international weeks were generated by interpolation and ages were grouped in ten–year width bins.

The 52–week (364–day) accumulated number of weeks for all–cause deaths and COVID–19 were computed from these tables. Results were divided by population numbers.

For every region and at week 08 age specific accumulated year-on-year mortality rates from 2002 to 2020 were fit to a Poisson generalized linear model with rates depending linearly on week number.^1^ Excess death rates were computed by the difference of observed 2021 values to predicted values. Same technique was performed for nationwide data.

Figure 1 displays the results for this ultimate set. Note that the youngest group age is not shown.

**FIG. 1.**
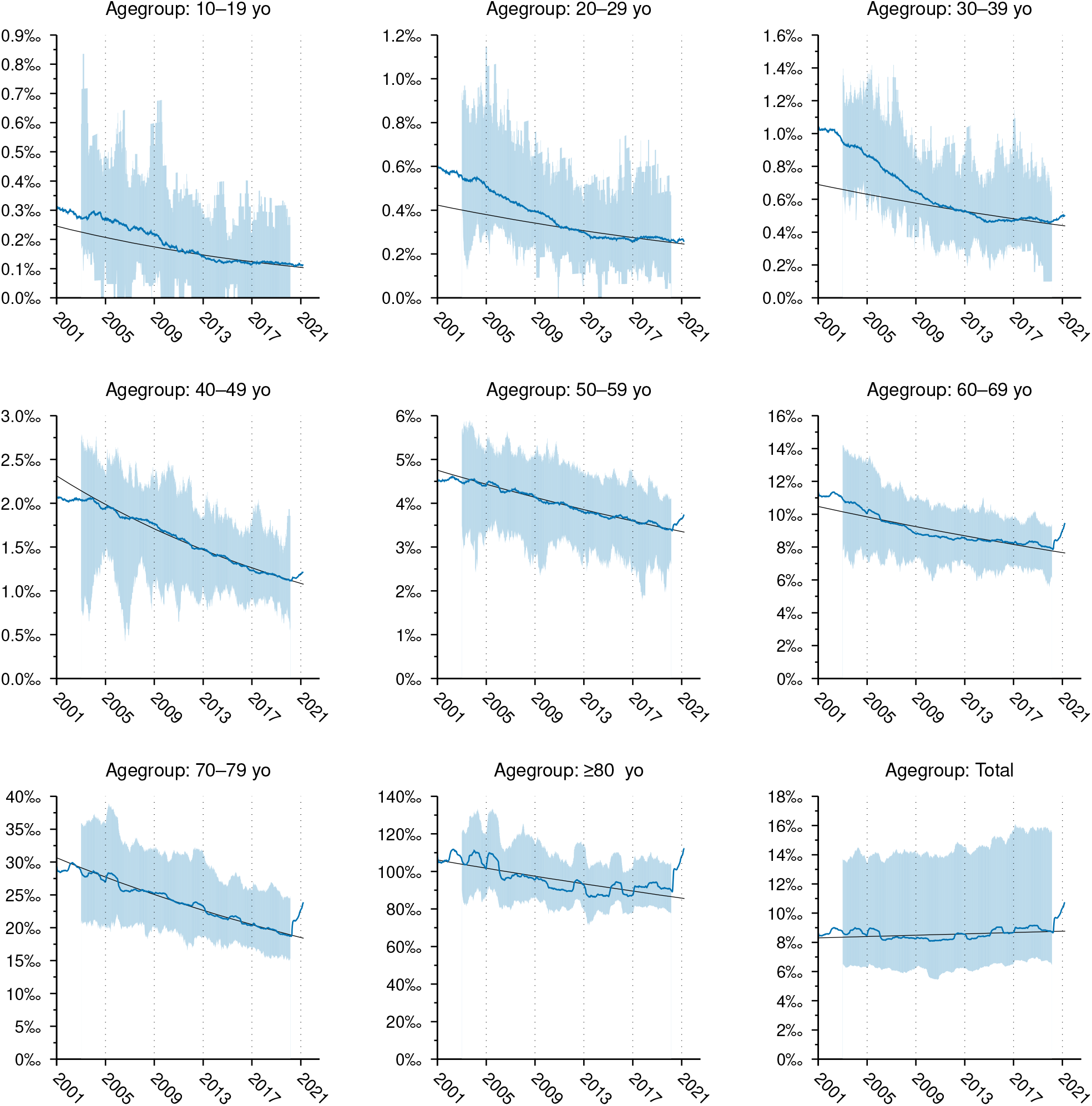
All–cause 52–week death rates in Spain for AGEGROUP10 (blue) and generalized linear Poisson model evaluated for weeks 09 from 2002 to 2020 (black). Excess death rates are computed by substracting the model (black) to the observations (blue). Shade displays the range where NUTS3 regional values can be found. For the younger age groups the model considered only from 2009 to 2020. The excursion following the pandemic is clearly observed in older age groups.

## 3. RESULTS AND DISCUSSION

Figure 2 shows a collection of nine scatter plots where all–cause excess death rates in 52 weeks (outcome, *y*) are tested against COVID–19 death rates (predictor, *x*). Each point represent one of the 52 Spanish NUTS3 regions; points symbols denote the same NUTS1 (supra-region) which are ES1 (North West), ES2 (North), ES3 (Madrid), ES4 (Center), ES5 (East), ES6 (South) and ES7 (Canary Islands). Solid lines show the predicted linear behaviour for every age group. Broken lines highlight the identity *y* = *x*.

**FIG. 2.**
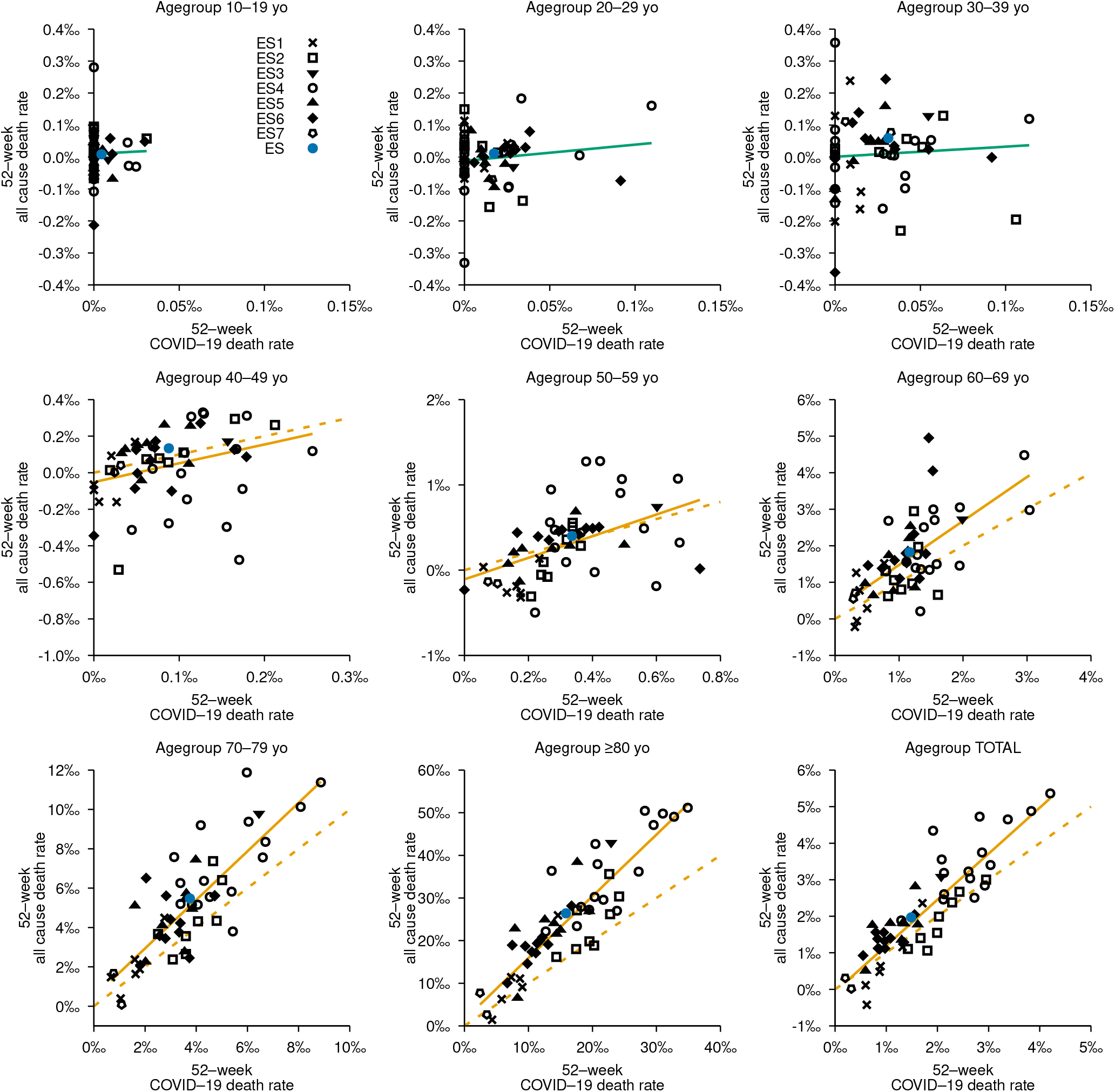
Age specific scatter plot for all–cause 52–week death rates (outcome, *y*) versus identified COVID–1952–week death rate (predictor, *x*) for Spanish NUTS3 regions (*N* = 52). Solid line shows the predicted behaviour. Broken line shows the hypothesis outcome equals predictor (*y* = *x*). Lines are displayed in orange ink if the null hypothesis does not sustain; else in light green. The youngest age group is not shown. NUTS3 regions sharing NUTS1 region display the same point symbol, see upper left panel. Nationwide values are displayed in blue ink.

Table 1 lists results for the bivariate analysis. This tests the hypothesis outcome and predictor are correlated. Results shows this is only statistically significant at the standard level for population aged 40 years or more (*p−* val *<* 0.05 in which case the row is shaded). Below this threshold all–cause excess death rates and COVID–19 deaths rates are low and the null hypothesis “outcome and predictor do not correlate” sustains.

**TABLE 1.** Age specific bivariate analysis for all–cause excess death rate (outcome, *y*) and identified COVID–19 death rate (predictor, *x*) for Spanish NUTS3 regions (*N* = 52). Slopes punctuate the COVID–19 undercount. The intercepts are given per one thousand population. When the null hypothesis “all–cause excess death rates do not depend on COVID–19 death rates” does not sustain at the standard level of significance the row is shaded in light gray, highlighting a correlation between COVID–19 deaths and excess deaths.

| Age group | Slope | | Intercept | | $p$-val | Pearson's $R^2$ |
| --- | --- | --- | --- | --- | --- | --- |
|  | Value | 95 % C.I. | Value | 95 % C.I. |  |  |
| 00–09 yo | 0.92 | [−0.30, 2.14] | −0.05 | [−0.09, 0.00] | 0.14 | 0.04 |
| 10–19 yo | 0.28 | [−2.15, 2.71] | 0.01 | [−0.01, 0.03] | 0.82 | 0.00 |
| 20–29 yo | 0.51 | [−0.55, 1.57] | −0.01 | [−0.04, 0.02] | 0.34 | 0.02 |
| 30–39 yo | 0.32 | [−1.01, 1.65] | 0.00 | [−0.05, 0.05] | 0.63 | 0.00 |
| 40–49 yo | 1.03 | [0.13, 1.93] | −0.05 | [−0.15, 0.04] | 0.03 | 0.10 |
| 50–59 yo | 1.27 | [0.64, 1.90] | −0.11 | [−0.33, 0.11] | $1.9 \times 10^{-4}$ | 0.25 |
| 60–69 yo | 1.20 | [0.80, 1.61] | 0.27 | [−0.24, 0.79] | $2.5 \times 10^{-7}$ | 0.42 |
| 70–79 yo | 1.23 | [0.98, 1.49] | 0.47 | [−0.57, 1.51] | $3.3 \times 10^{-13}$ | 0.66 |
| ≥ 80 yo | 1.45 | [1.24, 1.66] | 1.45 | [−2.33, 5.22] | 0.0 | 0.79 |
| All ages | 1.26 | [1.07, 1.44] | −0.07 | [−0.42, 0.29] | 0.0 | 0.79 |

The slope in Table 1 is the COVID–19 undercount. In contrast, regional undercount value can be depicted from the the slope of the straight line joining a data point and the origin, not shown in the plot.

However the statistical significance of the undercount must be further analyzed. The first point already mentioned above 40 years old COVID–19 deaths correlate to all–death, meaning the pandemic and all–cause deaths are correlated. It is worth to mention outcome and predictor come from independent reports.

The second thing to assess is to what extend the relative undercount is significant. For this to be analyzed one should either test the significance of the slope against the value one —*y* = *x*, which would mean correlation with no undercount— or, and this is the same, do a bivariate analysis where the outcome is now non-COVID–19 death rate defined as the difference from all–cause deaths to COVID–19 deaths. In other words the absolute undercount tested from *y − x* versus *x*.

Table 1 shows that unit slope is covered in the 95 % confidence interval for every age group below 80 years old, meaning that statistically speaking undercount is only observed in this age group. Given the fact that this eldest age group contributes the most to the total outcome, the significance also propagates to the total age count. Table 2 lists results for the bivariate analysis and Figure 3 shows the scatter plots. Regardless of the statistical significance the predicted values of non–COVID–19 death rate slope for the eldest age group doubles those of younger groups.

**TABLE 2.** Age specific bivariate analysis for non–COVID–19 52–week excess death rate (outcome) and identified 52–week COVID–19 death rate —*y − x* against *x* as of Table 1— in Spanish NUTS3 regions (*N* = 52). The intercepts are given per one thousand population. When the null hypothesis “non–COVID–19 excess death rates do not depend on COVID–19 death rates” does not sustain at the standard level of significance the row is shaded in light gray, highlighting significant non–excess deaths.

| Age group | Slope | | Intercept | | $p$-val | Pearson's $R^2$ |
| --- | --- | --- | --- | --- | --- | --- |
|  | Value | 95 % C.I. | Value | 95 % C.I. |  |  |
| 00–09 yo | −0.08 | [−1.30, 1.14] | −0.05 | [−0.09, 0.00] | 0.90 | 0.00 |
| 10–19 yo | −0.72 | [−3.15, 1.71] | 0.01 | [−0.01, 0.03] | 0.56 | 0.01 |
| 20–29 yo | −0.49 | [−1.55, 0.57] | −0.01 | [−0.04, 0.02] | 0.35 | 0.02 |
| 30–39 yo | −0.68 | [−2.01, 0.65] | 0.00 | [−0.05, 0.05] | 0.31 | 0.02 |
| 40–49 yo | 0.03 | [−0.87, 0.93] | −0.05 | [−0.15, 0.04] | 0.94 | 0.00 |
| 50–59 yo | 0.27 | [−0.36, 0.90] | −0.11 | [−0.33, 0.11] | 0.40 | 0.01 |
| 60–69 yo | 0.20 | [−0.20, 0.61] | 0.27 | [−0.24, 0.79] | 0.32 | 0.02 |
| 70–79 yo | 0.23 | [−0.02, 0.49] | 0.47 | [−0.57, 1.51] | 0.07 | 0.06 |
| ≥ 80 yo | 0.45 | [0.24, 0.66] | 1.45 | [−2.33, 5.22] | $9.2 \times 10^{-5}$ | 0.27 |
| All ages | 0.26 | [0.07, 0.44] | −0.07 | [−0.42, 0.29] | $6.9 \times 10^{-3}$ | 0.14 |

**FIG. 3.**
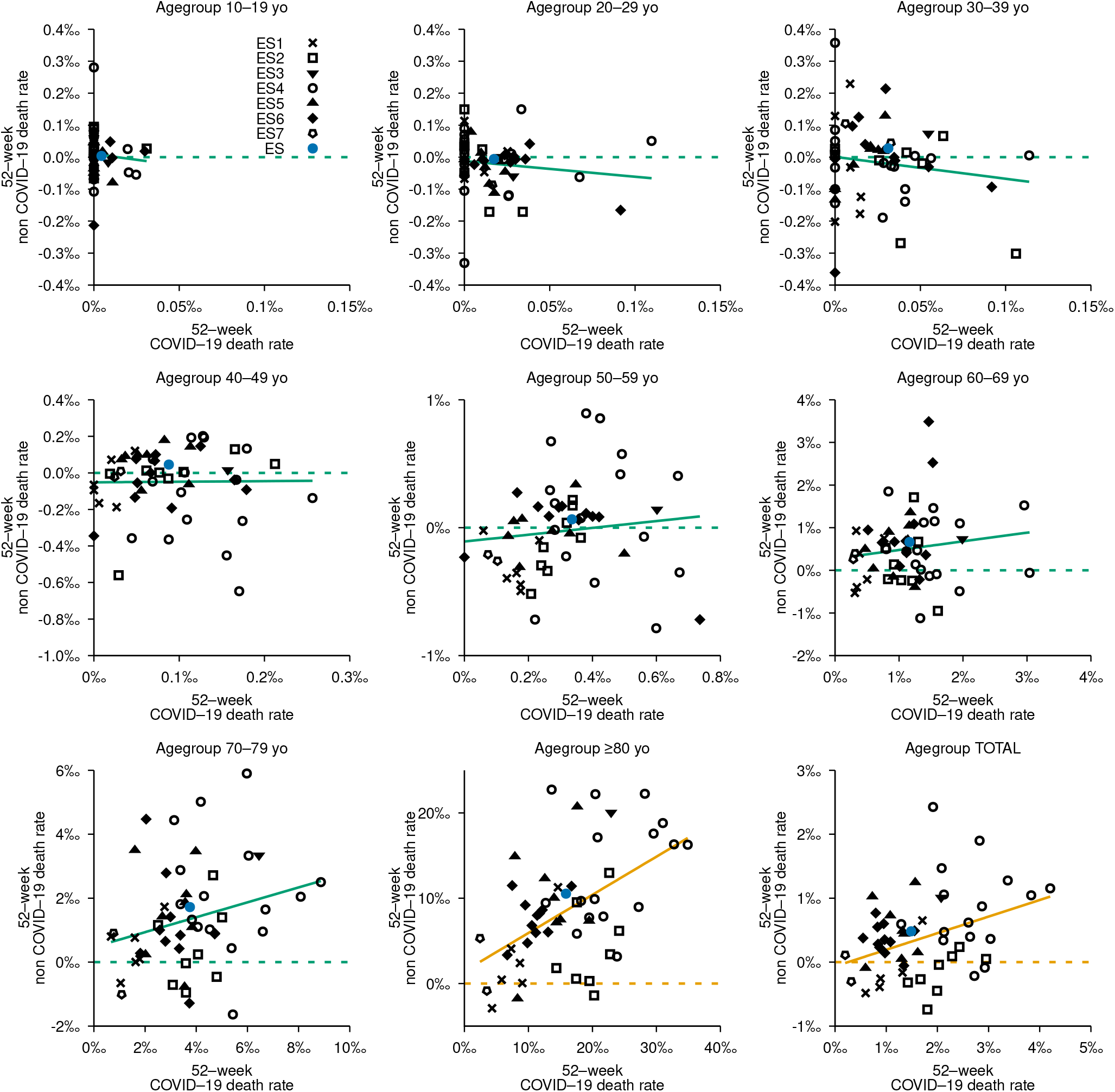
Age specific scatter plots of non–COVID–19 52–week death rates (outcome) versus identified COVID–1952–week death rate (predictor) —*y− x* versus *x* as of Figure 2— for Spanish NUTS3 regions (*N* = 52). Solid line shows the predicted behaviour. Broken line shows the null hypothesis of no non–COVID–19 excesses (*y* = 0). Lines are displayed in orange ink if the null hypothesis does not sustain; else in light green. The youngest age group is not shown. NUTS3 regions sharing NUTS1 region display the same point symbol, see upper left panel. Nationwide values are displayed in blue ink.

Table 1 and Table 2 predicts all–cause deaths and non–COVID–19 deaths from observed COVID–19 deaths. For the nationwide and all ages statistics, COVID–19 death toll is *x* = 70 366 and *y* = 93 250 (all–cause deaths) thus making *y− x* = 22 888 non–COVID–19 deaths. Table 1 predicts 88 661 all–cause deaths for the given *x* and Table 2 predicts 18 295 non–COVID–19 deaths for the given *x*. For population ageing 80 or elder, *x* = 45 335 and *y* = 75 445 or *y−x* = 30 110. Table 1 *x* predicts 65 735 all–cause deaths and Table 2 predicts 20 400 non–COVID–19 deaths, larger than the prediction for all ages. This exemplifies that the burden of the undercount may rest on the eldest age group.

In the early stages of the outbreak institutionalized elder population was heavily impacted by the disease. The flood of infections and deaths prevented authorities from reporting accurate numbers which only few months later surfaces now and then[6]^2^.

Results from this analysis agrees with this history. Indeed for population aged 80 or elder the highest non–COVID–19 death rates are observed (see Figure 3) in many ES4 provinces (circles), Madrid (ES3, down triangle) and in Barcelona (ES5, up triangle) which agrees with press reports on COVID–19 related deaths in Spanish long-term care facilities (see footnote). Undercount is heavily observed in the eldest age group which is likely prone to have higher percentage of institutionalized population. This remark may even add to the fact that this population is potentially proner to be impacted by the stress of the public health system resulting in a greater mortality. Further analysis should enrich the causes of these terrible losses.

## 4. CONCLUSIONS

The correlation between all–deaths and COVID–19 deaths in Spain disaggregated by NUTS3 region and AGEGRP10 has been analyzed.

Results show significant correlation for age 40 yo or elder, meaning the impact of the diseases can be statistically traced up to this threshold.

Statistically significant non–COVID–19 excess deaths were identified for population ages 80 yo or elder as a result of the impact of the disease in institutionalized population and the stress of the public health system.

## Data Availability

All data come from public records. Links are provided in the manuscript.

## ACKNOWLEDGMENTS

JMMO hearthfully thanks Eurostat, the Instituto Nacional de Estadística and the Centro Nacional de Epidemiología for making publicly available their datasets.

This work was performed using free software running on xubuntu 20.04.1LTS. Data sets have been imported into GNU octave-5.2.0 (https://www.gnu.org/software/octave/). Generalized linear Poisson model was performed on R https://www.r-project.org/. Pictures were developed thanks to gnuplot-5.2.8 (http://www.gnuplot.info/). The manuscript was typeset in GNU emacs-25.2.2 (https://www.gnu.org/software/emacs/) assisted by AUCTEX (https://www.gnu.org/software/auctex/). Data in pictures and in tables have been exported directly from octave.

This project started on March, 15 2021.

## Footnotes

1 Week numbers for week 09 starts at 9 for week 09, 2020, and are sequentially spaced by 52 weeks —week 09, 2021 is 61— in regular years and by 53 weeks in non–regular week years (2004, 2009, 2015 and 2020 in the record).

2 See also https://elpais.com/sociedad/2021-03-02/en-espana-han-muerto-29408-mayores-que-vivian-en-residencias-desde-el-inicio-dhtml, dated March 2, 2021 and were Spanish Government reports 30 000 COVID–19 related deaths on Spanish care facilities. However it is hard to say which fraction of them were previously accounted in official COVID–19 records.

